# Stress symptoms and reactions to COVID-19: A multinational survey from six Asian regions

**DOI:** 10.1101/2020.08.24.20179762

**Authors:** Ricci P.H. Yue, Edmund W. Cheng, Nick H.K. Or, Samson W.H. Yuen

**Author notes:** Corresponding author. E-mail address [R.P.H. Yue].

## Abstract

People may experience a heightened level of stress reactions during a pandemic event and in an isolated social environment. A multi-national survey about such mental health information about COVID-19 was conducted in May 2020 across six Asian regions: Hong Kong, Japan, Singapore, South Korea, Taiwan and Thailand. Data were collected from a population representative sample of 11,895 adults on their stress symptoms and reactions to COVID-19 and the related public health measures. 59.1% of the respondents showed at least one substantial stress symptom. The situation was particularly worrying in South Korea, where 75% of the respondents reported substantial stress symptoms. Respondents who were young, educated, lived in an urban area, had a high socioeconomic status, had a history of chronic illness or mental illness, or who lived with a pregnant woman, elderly or children were most vulnerable to stress during the pandemic. Stress reactions showed a positive relationship with the amount of time spent following news about the COVID-19 outbreak. Asian adults coped with their stress by preparing safety equipment and extra daily commodities (62.4%) and by following government-issued public health measures (60.1%). Most Asian (71%) also frequently checked on the safety of family members and friends to keep each other safe. The COVID-19 pandemic and the associated stringent public health measures have largely increased the prevalence of substantial stress symptoms across multiple Asian regions. Governments should prepare this mental health pandemic and the associated social repercussions.

## Introduction

The effects of the COVID-19 pandemic are severe, not only because of the lives it has cost, but also because of the strict public health measures implemented in most parts of the world. Drastic, immediate and austere public health measures ^1,2^ have been enacted in the hope of slowing the pandemic as much as possible. Yet, with the growing magnitude of the traumatic pandemic and the lengthening of tight public health measures, many people are developing symptoms of stress ^3,4^. The mental health crisis is undoubtedly worrying for most in Asia, where the novel coronavirus was first discovered and spread.

Few empirical studies have focused on stress symptoms among the public during a pandemic. The most relevant works, from Southeast Asia during the SARS and H1NI outbreaks ^5,6^, examined the public’s behavioural responses rather than stress symptoms. Recent studies on stress symptoms due to COVID-19 have focused mainly on either medical workers ^7,8^ or survivors ^9^, and others were confined to a local scale^10^. The absence of specific knowledge on population-wide stress symptoms during a catastrophic pandemic is troubling for the upcoming mental health crisis, which most assuredly will occur.

This study addressed three key issues. First, we surveyed nationally representative samples to determine the patterns and magnitudes of stress symptoms across six Asian regions since the early wave of COVID-19. Second, we characterised the groups that are vulnerable to stress during this pandemic according to variations in the full sample. Third, we asked about coping behaviours when faced with stress and the other reactions to COVID-19 and tightened public health measures. Thus, this study fills a gap in the literature by presenting a cross-national survey on the mental health of citizens during the COVID-19 pandemic.

## Methods

### Study design and data collection

We used a probability-based Internet panel in six Asian regions: Hong Kong, Japan, Singapore, South Korea, Taiwan and Thailand. Data were collected from May 11 to 26, when these six regions were entering the fifth month of the pandemic. The six sampled regions were selected for several reasons. First, they are regions that neighbour China, where the pandemic began. Second, they were all hit by the pandemic in the first wave of the disease in late January. Third, the scale of the outbreak differed among them, thus offering a comprehensive picture of stress symptoms over different pandemic settings.

The 10-minute survey was administered by Dynata, a global online survey research agency. The survey included questions on demographic information, mental health and chronic illness records for the past 12 months, household member information, amount of time spent following news about COVID-19, selected stress symptoms, coping behaviours and voluntary activities regarding COVID-19. The sample was selected according to the geographical and demographic features of the population based on the latest census data from each region. Quota sampling allowed us to assemble a pool of population representative responses. About 2000 responses were collected from each chosen region, for a total of 11,895 representative samples.

### Questionnaire design and key measurements

The Posttraumatic Stress Disorder Checklist^11^ was included in the questionnaire design to measure stress symptoms. Five stress symptoms were selected from the checklist based on their relevance to COVID-19 and from previous stress symptom reports of traumatised adults from the September 11 attacks^12^. The contexts of the stress symptom were modified from the COVID Stress Scale developed by Taylor, Landry ^13^. ‘Substantial stress symptoms’ were defined as the three highest options on a 7-point Likert scale^14^. Respondents who reported one or more substantial stress symptoms were considered to have a ‘substantial stress condition’. The scores from all five stress symptom questions were added to form a ‘generalised stress level’ score for further statistical analysis. The questions related to stress coping behaviour were treated in the same way as the stress symptom data. The total score from all stress coping behaviour-related questions was added to form a ‘stress coping actions’ score.

To assess the coping reactions and potential predictors of the stress symptoms, we developed other questionnaire items with reference to existing studies, advice from the World Health Organization and media reports. We considered the respondents’ mental health records because they are the key determinants of stress reactions^15^, and their chronic illness records because they contain risk factors for severe COVID-19 cases and deaths^16^. To assess the impact of having a vulnerable household member, we asked whether the respondent lived with a pregnant woman, an adult older than 65 years or a child younger than 12 years. The questionnaire also measured the average time the respondent spent viewing information about COVID-19 each day. The time frame for all questions was set to the COVID-19 outbreak in each surveyed region.

### Statistical analysis

Data were analysed with Stata software and with linear regression to outline the association between the dependent variables and the predictor variables. Our analysis was conducted at the overall response level and at the regional level. We report the alpha level at 90%, 95%, 99% and 99.5% confidence intervals for statistical significance. For transformed variables such as the ‘generalised stress level’ and ‘stress coping action’, we measured internal consistency using Cronbach’s alpha for the set of items before collapsing the responses into a summed score.

### Results

As high as 59.1% of the 11,895 respondents in the six Asian regions surveyed had experienced at least one of the substantial stress symptoms since the COVID-19 outbreak (Table 1). The region with the most people with a substantial stress condition was South Korea (N=2030) (75.0%), followed by Hong Kong (60.3%) (N=1846) and Thailand (59.9%) (N=2021), and the region with the fewest was Taiwan (45.3%) (N=2025). The most common substantial stress symptom (46.9%) was ‘stressed about leaving home’, which was reported by 63.4% of the South Korean respondents. In the six Asian regions, 26.1% to 44.4% of adults reported having repetitive and disturbing thoughts or dreams about what was happening in the pandemic; 22.7% to 41.2% and 20.7% to 34.0% of the respondents within the surveyed regions had trouble concentrating and trouble falling or staying asleep, respectively; and 20.4% to 43.1% of the people felt irritable or had outbursts of anger.

**Table 1.**
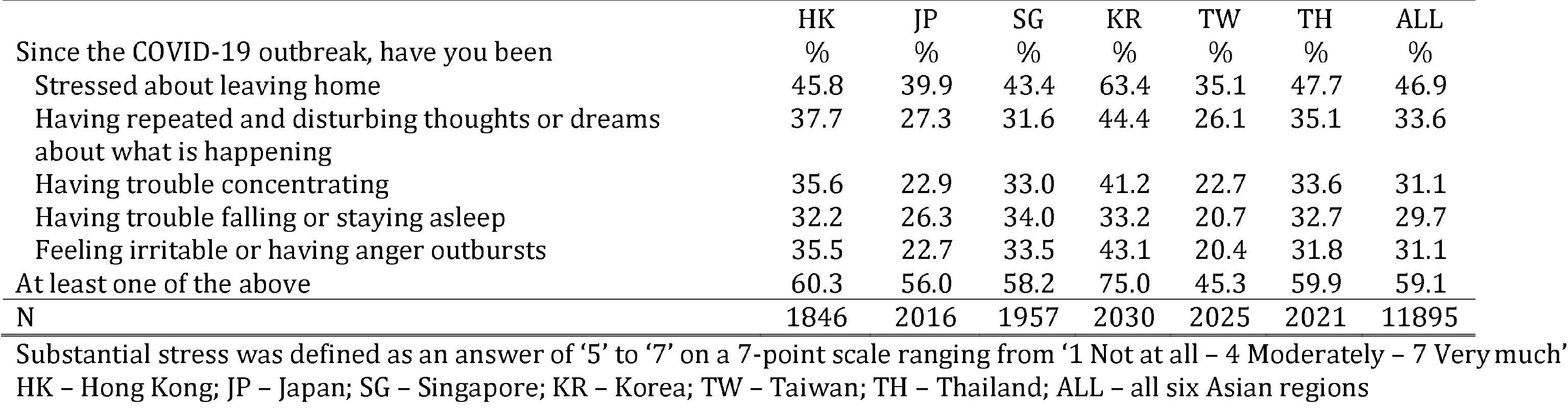
Level of substantial stress in six Asian regions during COVID-19 outbreak

|  |  |  |  |  |  |  |  |
| --- | --- | --- | --- | --- | --- | --- | --- |
|  | HK | JP | SG | KR | TW | TH | ALL |
| Since the COVID-19 outbreak, have you been | % | % | % | % | % | % | % |
| Stressed about leaving home | 45.8 | 39.9 | 43.4 | 63.4 | 35.1 | 47.7 | 46.9 |
| Having repeated and disturbing thoughts or dreams about what is happening | 37.7 | 27.3 | 31.6 | 44.4 | 26.1 | 35.1 | 33.6 |
| Having trouble concentrating | 35.6 | 22.9 | 33.0 | 41.2 | 22.7 | 33.6 | 31.1 |
| Having trouble falling or staying asleep | 32.2 | 26.3 | 34.0 | 33.2 | 20.7 | 32.7 | 29.7 |
| Feeling irritable or having anger outbursts | 35.5 | 22.7 | 33.5 | 43.1 | 20.4 | 31.8 | 31.1 |
| At least one of the above | 60.3 | 56.0 | 58.2 | 75.0 | 45.3 | 59.9 | 59.1 |
| N | 1846 | 2016 | 1957 | 2030 | 2025 | 2021 | 11895 |

Substantial stress was defined as an answer of '5' to '7' on a 7-point scale ranging from '1 Not at all – 4 Moderately – 7 Very much'
HK – Hong Kong; JP – Japan; SG – Singapore; KR – Korea; TW – Taiwan; TH – Thailand; ALL – all six Asian regions

As for the COVID-19 information viewing pattern, people in the six regions spent an average of 3.3 hours (SD=4.3 hours) each day reading news about the coronavirus outbreak. Thailand stood out as the country in which the most attention was paid to COVID-19-related news (average=5.7 hours; SD=5.7 hours). The average time spent viewing news on the COVID-19 pandemic was 2.6 hours (SD=3.5 hours) in Hong Kong, 2.6 hours (SD=3.3 hours) in Japan, 3.3 hours (SD=4.2 hours) in Singapore, 2.9 hours (SD=3.9 hours) in South Korea and 2.6 hours (SD=3.6 hours) in Taiwan.

We characterised the vulnerable community by predicting the risk factors for a generalised stress condition (alpha=0.916; Table S6). In general, people who are younger (P<0.005), more educated (P<0.005), lived in an urban environment (P<0.005) and had a high socio-economic status (P<0.005) were statistically significantly more prone to have a stress condition (Table 2). Patients with a history of chronic illness (P<0.005) and those with mental or emotional history during the year before the pandemic (P<0.005) were more susceptible to stress. Family members who lived with a pregnant woman or an older adult over 65 years (P<0.01) and children younger than 12 years (P<0.005) were also more vulnerable to a stress condition. The time spent each day following news about the COVID-19 outbreak was positively correlated with generalised stress level (P<0.005). In the regional-scale analysis, the risk factors for a stress condition varied among the six regions (Table 3). However, having mental or emotional problems before the outbreak, having children younger than 12 years in the household and the total time spent viewing COVID-19 information remained as statistically significant predictors of a stress condition in all six regional settings. The regressions were repeated for each type of stress symptom for both the full dataset and for each region; age, a history of mental or emotional problems, having children younger than 12 years in the household and the total time spent viewing COVID-19 information were predictors of all symptoms in most cases (Table S1-S5).

**Table 2.**
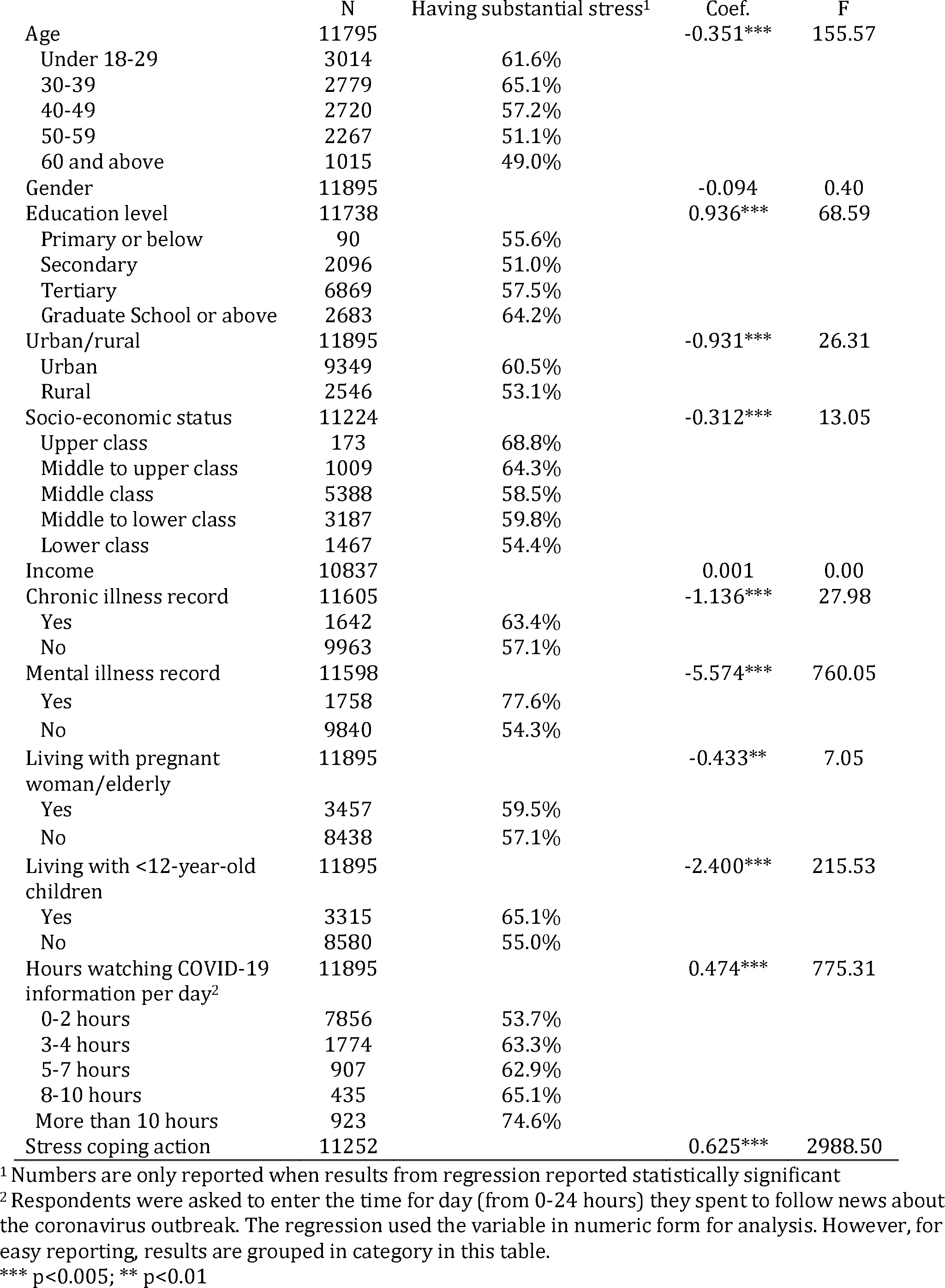
Risk factors for generalised stress level in Asia

**Table 3.**
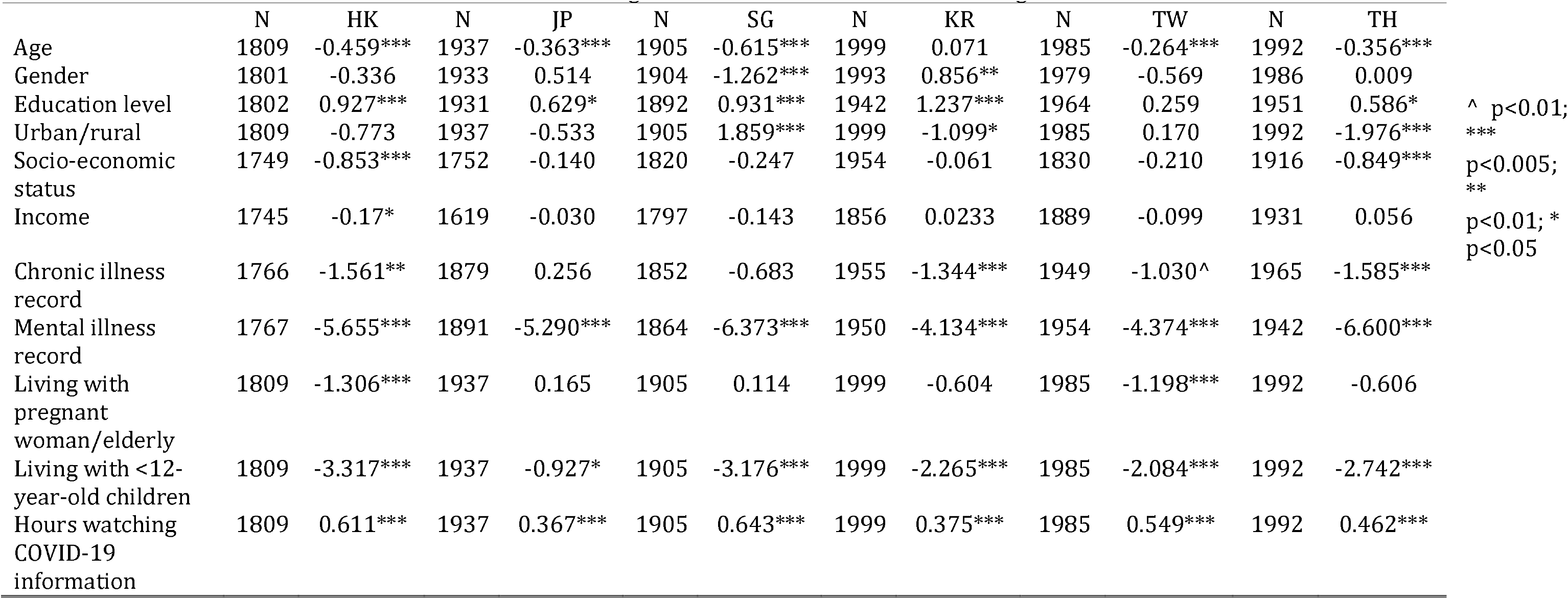
Risk factors for generalised stress level in six Asian regions

|  | N | HK | N | JP | N | SG | N | KR | N | TW | N | TH |
| --- | --- | --- | --- | --- | --- | --- | --- | --- | --- | --- | --- | --- |
| Age | 1809 | -0.459*** | 1937 | -0.363*** | 1905 | -0.615*** | 1999 | 0.071 | 1985 | -0.264*** | 1992 | -0.356*** |
| Gender | 1801 | -0.336 | 1933 | 0.514 | 1904 | -1.262*** | 1993 | 0.856** | 1979 | -0.569 | 1986 | 0.009 |
| Education level | 1802 | 0.927*** | 1931 | 0.629* | 1892 | 0.931*** | 1942 | 1.237*** | 1964 | 0.259 | 1951 | 0.586* |
| Urban/rural | 1809 | -0.773 | 1937 | -0.533 | 1905 | 1.859*** | 1999 | -1.099* | 1985 | 0.170 | 1992 | -1.976*** |
| Socio-economic status | 1749 | -0.853*** | 1752 | -0.140 | 1820 | -0.247 | 1954 | -0.061 | 1830 | -0.210 | 1916 | -0.849*** |
| Income | 1745 | -0.17* | 1619 | -0.030 | 1797 | -0.143 | 1856 | 0.0233 | 1889 | -0.099 | 1931 | 0.056 |
| Chronic illness record | 1766 | -1.561** | 1879 | 0.256 | 1852 | -0.683 | 1955 | -1.344*** | 1949 | -1.030^ | 1965 | -1.585*** |
| Mental illness record | 1767 | -5.655*** | 1891 | -5.290*** | 1864 | -6.373*** | 1950 | -4.134*** | 1954 | -4.374*** | 1942 | -6.600*** |
| Living with pregnant woman/elderly | 1809 | -1.306*** | 1937 | 0.165 | 1905 | 0.114 | 1999 | -0.604 | 1985 | -1.198*** | 1992 | -0.606 |
| Living with <12-year-old children | 1809 | -3.317*** | 1937 | -0.927* | 1905 | -3.176*** | 1999 | -2.265*** | 1985 | -2.084*** | 1992 | -2.742*** |
| Hours watching COVID-19 information | 1809 | 0.611*** | 1937 | 0.367*** | 1905 | 0.643*** | 1999 | 0.375*** | 1985 | 0.549*** | 1992 | 0.462*** |

^ p<0.01; \*\*\*p<0.005; \*\*p<0.01; \* p<0.05

To cope with stress, 62.4% of the Asian respondents showed a substantial tendency to prepare safety equipment and extra daily commodities (Table 4); 60.1% followed government-issued public health-related guidelines and orders to make themselves feel safer as a substantial stress coping behaviour; and 57.3% and 42.4% of the respondents, respectively, improved their lifestyles to stay healthier and talked with someone about their thoughts and feelings to cope with stress. The figures differed among the six Asian regions. Notably, Japan had a lower level of substantial stress coping behaviour than the other Asian regions. Stress coping actions (alpha=0.764; Table S6) showed a statistically significant association with the generalised stress level (P<0.005; Table 2).

**Table 4.** Level of substantial stress coping behaviour in six Asian regions during COVID-19 outbreak

|  | HK | JP | SG | KR | TW | TH | ALL |
| --- | --- | --- | --- | --- | --- | --- | --- |
| How do you cope with your emotions resulting from the pandemic | % | % | % | % | % | % | % |
| I talk with someone about my thoughts and feelings. | 46.7 | 27.5 | 40.0 | 52.8 | 39.9 | 47.6 | 42.4 |
| I improve my lifestyle to stay healthier . | 64.2 | 34.3 | 55.7 | 64.8 | 54.5 | 71.3 | 57.3 |
| I prepare safety equipment (i.e., facemasks) and extra daily commodities (i.e., food, gas, cash). | 75.5 | 33.1 | 58.2 | 67.1 | 66.3 | 75.6 | 62.4 |
| I turn to prayers and my religion. | 31.3 | 11.6 | 40.3 | 26.2 | 22.9 | 41.6 | 28.9 |
| I tend to follow the public health-related guidelines and orders issued by the government to make myself feel safer, | 48.9 | 33.2 | 71.5 | 69.5 | 60.1 | 76.2 | 60.1 |
| N | 1846 | 2016 | 1957 | 2030 | 2025 | 2021 | 11895 |

Substantial stress coping behaviour was defined as an answer of '5' to '7' on a 7-point scale ranging from '1. Not at all – 4. Moderately – 7 Very much'
HK – Hong Kong; JP – Japan; SG – Singapore; KR – Korea; TW – Taiwan; TH – Thailand; ALL – all six Asian regions

Table 5 summarises the voluntary responses in Asia during the COVID-19 outbreak. Substantial numbers of respondents checked on the safety of family members and friends to a moderate or frequent extent (71.0%), stocked supplies (41.8%), donated money (32.2%) and participated in voluntary activities (29.5%). The statistics for each type of voluntary response varied within a narrow margin (5.3% to 16.0%) within Hong Kong, Singapore, South Korea, Taiwan and Thailand. However, the figures were particularly low in Japan.

**Table 5.** Level of voluntary response in six Asian regions during COVID-19 outbreak

|  | HK |  | JP |  | SG |  | KR |  | TW |  | TH |  | ALL |  |
| --- | --- | --- | --- | --- | --- | --- | --- | --- | --- | --- | --- | --- | --- | --- |
| How frequently have you engaged in the following actions | % |  | % |  | % |  | % |  | % |  | % |  | % |  |
|  | Yes | Moderately to frequently | Yes | Moderately to frequently | Yes | Moderately to frequently | Yes | Moderately to frequently | Yes | Moderately to frequently | Yes | Moderately to frequently | Yes | Moderately to frequently |
| Donated money, masks or other supplies to those who might need them | 85.3 | 39.6 | 41.9 | 12.7 | 71.1 | 29.8 | 71.5 | 31.1 | 86.7 | 37.6 | 88.2 | 42.9 | 73.9 | 32.2 |
| Got hold of any extra food, gas, cash, or other supplies you might need | 93.0 | 55.8 | 71.0 | 21.8 | 84.0 | 39.8 | 82.0 | 42.2 | 88.3 | 43.0 | 90.7 | 49.3 | 84.7 | 41.8 |
| Checked the safety of family members and friends | 98.0 | 78.1 | 86.6 | 45.1 | 97.4 | 72.3 | 97.6 | 77.2 | 98.1 | 76.5 | 98.2 | 77.6 | 95.8 | 71.0 |
| Participated in voluntary activities to help those who are vulnerable to the virus | 80.3 | 35.9 | 30.2 | 9.6 | 66.3 | 29.2 | 75.8 | 36.8 | 83.9 | 32.4 | 78.5 | 36.6 | 69.0 | 29.5 |
| N | 1846 |  | 2016 |  | 1957 |  | 2030 |  | 2025 |  | 2021 |  | 11895 |  |

HK – Hong Kong; JP – Japan; SG – Singapore; KR – Korea; TW – Taiwan; TH – Thailand; ALL – all six Asian countries

## Discussion

The results of our cross-national survey show that a remarkably high number of people in all six Asian regions experienced symptoms of stress during the COVID-19 outbreak. Many respondents (59.1%) had been affected substantially by at least one stress symptom since the pandemic outbreak. The situation was particularly worrying in South Korea, where 75.0% of the respondents showed at least one substantial stress symptom. The findings indicate that COVID-19 has extended from a pandemic into a mental well-being crisis in Asia. The prevalent signs of post-traumatic stress – stress, disturbed thoughts or dreams, sleeping problems, trouble concentrating and anger outbursts – pinpoint early diagnosis of post-traumatic stress disorder in the context of a high hidden cost of strict public health measures and the widespread scale of the COVID-19 outbreak in the selected regions.

A few recent studies have also specified the psychological stress effects of the COVID-19 outbreak. An early study from China conducted in late January reported that 53.8% of the public considered the COVID-19 outbreak as having a moderate or severe psychological impact, and 8.1% of the respondents reported moderate to severe stress levels^4^. Another study from China in mid-February indicated that the trauma level and psychological stress level of the lay public were higher than those of the nurses at the front-line in hospitals^17^. A recent report with data collected from April to May implied that 25.4% of people in Hong Kong reported a deterioration in their mental health during the pandemic^10^. Our results indicate that psychological stress in major Asian regions may have shown exponential growth between February and May. This signifies that the combination of the pandemic itself and the stringent public health measures and social distancing might have caused the mass spread of stress and panic. A moderate level of stress and panic promotes alertness to developments and obedience to COVID-19–related protective habits such as wearing masks and washing hands^18^. However, a prolonged state of traumatic stress may not help in dealing with the long-term threat.

We also compared our stress figures with those from previous disasters. Data from a few days after the September 11 attacks in 2001 suggested that the proportion of the US population that experienced substantial stress symptoms was only half the proportion of the Asian population during the COVID-19 outbreak^12^. Another study on the Oklahoma City bombing attack in 1995 indicated that the stress symptoms surveyed under our study were about half the level of those who survived the direct blast of that bombing event ^19^. Methodological discrepancies might have generated potential error in such comparisons, but our results indicate that by May 2020, COVID-19 had driven stress symptoms to a disastrous level among the Asian public.

In particular, our results indicate a surging risk of anger outbursts (31.1%) in underprivileged young urban families with children. Such frustration and irritation could easily fuel potential conflict. In Asian metropolitan cities such as Hong Kong, limited living space has been a challenge for families during the city lockdown. Isolation, paired with psychological stressors and an economic downturn, can combine to trigger domestic violence on an unprecedented scale^20,21^.

The risk of substantial stress symptoms varied according to the respondents’ demographic backgrounds. The catastrophe seems to have had a more pronounced stress effect on citizens who are young, educated, live in urban areas and have a high socio-economic status. Stress symptoms were also more prevalent among those who had a history of chronic illness or mental illness, lived with a pregnant woman, an adult older than 65 years or a child younger than 12 years. There are three possible reasons for such a pattern. First, young and educated individuals have been deluged with information and messages during this pandemic, leading to elevated stress. Second, when pension and housing wealth of urban richer groups were taking a big hit during the pandemic, in short run the immediate impact of the crisis might seem more devastating to the rich people in cities ^22^. Yet, other reports also suspected socially disadvantaged individuals should have a disproportionate likelihood of being affected by COVID-19 ^23^. Third, individuals with limited immune capacity, stress capacity or mobility were more likely to experience greater fear and extra stressors than the others ^24^. These predictors allowed us to identify vulnerable groups to formulate a strategic response plan to the public panic crisis during a pandemic.

Our survey also indicated that the level of stress was positively associated with the average number of hours a day spent viewing COVID-19-related information. Traditionally, the threat appraisal model holds that viewing information can be a positive coping mechanism for predicting outcomes in the face of risk and uncertainty ^25^. However, recent studies have noted that the vast amount of online information available in the digital era and the excessive use of social media during the pandemic could contribute to cyberchondria and information overload^26,27^, which could further generate confusion and stress.

The stress coping behaviour of the Asian respondents showed a statistically significant relationship with stress. They tended to prepare safety equipment and extra daily commodities, to adopt a healthier lifestyle or to follow government-issued public health guidelines and orders to cope with their stress. Taking such constructive actions suggests that Asian people try to reduce uncertainty to cope with stress and anxiety ^28^. The respondents also tended to show voluntary responses amidst the pandemic. Previous reports indicated that helping from the victims of a tragedy from distant is regarded as a collective social response in coping with the aftermath of trauma ^12,29^. A further study found that compassion was regarded as an active process through which people show kindness and support, which, in turn, can serve as an intervention for their depression or anxiety^30^.

This study has limitations. The study design did not incorporate a full assessment of post-traumatic stress disorder and thus may have underestimated the respondents’ level of stress or trauma. However, the strength of this study is the stress symptom data obtained from a vast population across six Asian regions differentially affected by the COVID-19 outbreak and with different responses to the pandemic. The data from our study contribute to a growing evidence base regarding the mental health crisis that is spreading along with the pandemic. Previous studies of disaster suggest that most stress symptoms fade over time, especially in those who experience the trauma indirectly. However, the current COVID-19 pandemic presented a chronic hazard with waves of overlapping outbreaks rather than a single acute one. Many people are unable to cope with uncertainty and stress for weeks and months. This can ultimately lead to exhaustion and depression, which could amplify the impact of COVID-19 and jeopardise decision-making when combating the pandemic. Although this is unlikely to anticipate the endpoint of the current COVID-19 pandemic, we call for additional early detection, intervention, public health resources and policy focus for those who are most vulnerable to stress and post-traumatic stress disorder. The global society must prepare for this storm.

### Contributors

RPHY, EWC, NHKO, and SWHY contributed to study concept and design. RPHY, EWC, NHKO, and SWHY contributed to acquisition of data. PRHY contributed to data analysis and initial drafting of the manuscript. All authors contributed to interpretation of data and critical revision of the report. RPHY, EWC, NHKO, and SWHY contributed to study supervision.

## Data Availability

Data is available upon requests to Dr. Ricci P.H. Yue at

## Acknowledgement

This study was supported by Knowledge Transfer Earmarked Fund from University Grants Commission (6250001). The funder had no role in the design of the study; the collection, analysis, and interpretation of the data; the writing of the manuscript; or the decision to submit the manuscript for publication.

